# Impact assessment of non-pharmaceutical interventions against COVID-19 and influenza in Hong Kong: an observational study

**DOI:** 10.1101/2020.03.12.20034660

**Authors:** Benjamin J. Cowling, Sheikh Taslim Ali, Tiffany W. Y. Ng, Tim K. Tsang, Julian C. M. Li, Min Whui Fong, Qiuyan Liao, Mike YW Kwan, So Lun Lee, Susan S. Chiu, Joseph T. Wu, Peng Wu, Gabriel M. Leung

## Abstract

**Background:** A range of public health measures have been implemented to delay and reduce local transmission of COVID-19 in Hong Kong, and there have been major changes in behaviours of the general public. We examined the effect of these interventions and behavioral changes on the incidence of COVID-19 as well as on influenza virus infections which may share some aspects of transmission dynamics with COVID-19.

**Methods:** We reviewed policy interventions and measured changes in population behaviours through two telephone surveys, on January 20-23 and February 11-14. We analysed data on laboratory-confirmed COVID-19 cases, influenza surveillance data in outpatients of all ages, and influenza hospitalisations in children. We estimated the daily effective reproduction number (*R*_*t*_), for COVID-19 and influenza A(H1N1).

**Findings:** COVID-19 transmissibility has remained at or below 1, indicating successful containment to date. Influenza transmission declined substantially after the implementation of social distancing measures and changes in population behaviours in late January, with a 44% (95% confidence interval, CI: 34% to 53%) reduction in transmissibility in the community, and a 33% (95% CI: 24% to 43%) reduction in transmissibility based on paediatric hospitalization rates. In the two surveys we estimated that 74.5% and 97.5% of the general adult population wore masks when going out, and 61.3% and 90.2% avoided going to crowded places, respectively.

**Implications:** Containment measures, social distancing measures and changes in population behaviour have successfully prevented spread of COVID-19. The social distancing measures and behavioural changes led to a substantial reduction in influenza transmission in early February 2020. However, it may be challenging to avoid fatigue and sustain these measures and population behaviours as COVID-19 continues to spread globally.

**Funding:** Health and Medical Research Fund, Hong Kong

## INTRODUCTION

The WHO-China Joint Mission on COVID-19 completed its work and tendered the final report on February 24, 2020.^1^ The mission concluded that there had been a substantial fall in reported case counts in mainland China since late January. These epidemic trends resulted from unprecedented, perhaps draconian, non-pharmaceutical interventions implemented, particularly since the *cordon sanitaire* was imposed on Wuhan and surrounding municipalities on January 23, 2020. The measures relied heavily on massive mobility restrictions within and between conurbations, universal fever screening in all settings, neighbourhood-based, household-focused social distancing that is enforced by large teams of community workers as well as pervasive deployment of AI-based, “big data” social media apps, amongst others.^2^ Whether some or all of these would be acceptable and feasible in settings outside of mainland China has been questioned.^3^

Hong Kong is a Special Administrative Region of China that operates with a large degree of autonomy. It is located outside the mainland on the southern coast of China, neighbouring Guangdong province. Guangdong has recorded the most confirmed COVID-19 cases (n=1356 as of 11 March) outside of Wuhan and its home province Hubei at the centre of the ongoing global outbreak. Having been one of the most heavily affected epicentres during the Severe Acute Respiratory Syndrome (SARS) epidemic in 2003, the community in Hong Kong have been particularly well prepared to respond to emerging infectious diseases. A range of public health measures have been implemented to delay and reduce local transmission of COVID-19, and there have been major changes in behaviours of the general public.

The containment measures used to control COVID-19 in Hong Kong include intense surveillance for infections not only in incoming travellers but also in the local community, with around 500 outpatients and 800 inpatients tested each day in early March (personal communication, GM Leung). Once cases are identified they are isolated until they recover and cease virus shedding. Their close contacts are traced (currently going back as far as two days before illness onset) and quarantined in special facilities, including holiday camps and newly constructed housing estates. Because not every infected person will be identified, containment measures only work if social distancing measures or behavioural changes also reduce “silent transmission” in the community as a whole.

Hong Kong is an ideal sentinel to study the impact of public health interventions and population behavioural changes that may be closer to what may plausibly be rolled out in resource-sufficient settings in the rest of the world. Here, we aimed to quantify the effect of containment measures on COVID-19. In addition, to determine whether social distancing and behavioural changes have been important in reducing silent transmission of COVID-19, we analysed data on influenza activity as a proxy to illustrate the potential changes in transmission of infection in line with the interventions implemented, assuming a similar mode and efficiency of spread between influenza and COVID-19. The specific objective of this study was to quantify population behavioural changes in Hong Kong during the outbreak of COVID-19, and to describe the impact of the behavioural changes and public health measures on COVID-19 transmission and influenza transmission in the community.

## METHODS

### Sources of Data

Data on laboratory-confirmed COVID-19 cases were obtained from the Centre for Health Protection who provide daily updates with individual case data on a dedicated webpage.^4^ We conducted two cross-sectional telephone surveys among the general adult population in Hong Kong. The first survey was conducted on January 20-23, and the second survey was conducted on February 11-14. Methods and survey instruments used were similar to those used to surveys during the SARS epidemic in 2003,^5,6^ the influenza A(H1N1)pdm09 pandemic in 2009,^7^ and the influenza A(H7N9) outbreak in China in 2013.^8^ Participants were recruited using random-digit dialling of both landline and mobile telephone numbers. Telephone numbers were randomly generated by a computer system. Calls were made during both working and non-working hours by trained interviewers to avoid over-representation of non-working groups. Respondents were required to be at least 18 years of age and able to speak Cantonese Chinese or English. Within each household, an eligible household member with the nearest birthday would be invited to participate in the survey. The same core questions on behavioural responses to the threat of COVID-19 were used in both surveys, with some supplementary questions asked in survey 1 or 2 but not both. Items included measures of risk perception, attitudes towards COVID-19, and behaviours taken against COVID-19 including hygiene, facemasks and reduction of social contacts. In the second survey, respondents who were parents of school-age children were asked to answer additional questions about social contact patterns of their children since schools were closed at the time of interview. Ethical approval for this study was obtained from the Institutional Review Board of the University of Hong Kong.

We obtained sentinel surveillance data on influenza-like illnesses (ILI) in a network of around 60 general outpatient clinics from the Centre for Health Protection. These include weekly reports of the proportion of outpatient consultations that were in patients with influenza-like illness defined as fever plus cough or sore throat. We obtained laboratory surveillance data from the Public Health Laboratory Services on influenza testing results on specimens from public hospitals and sentinel surveillance sites including the weekly number of specimens tested and the number testing positive for influenza by type/subtype. Data on the current population of Hong Kong by age and sex were obtained from the Census and Statistics Department of the Hong Kong Government. We obtained the daily hospitalization rates for influenza positive cases in Hong Kong, using the daily hospital admissions for influenza to the paediatric departments of two large hospitals in Hong Kong and the relevant catchment populations.^9^

### Statistical analysis

Means and proportions of survey responses were directly weighted by sex and age to the general population. Categorical variables with ordinal Likert-type response scales including risk perception and attitudes towards COVID-19 were first dichotomized either above or below a threshold.

We estimated changes in COVID-19 transmissibility over time via the effective reproduction number, R_t_, which represents the mean number of secondary infections that result from a primary case of infection at time t. Values of R_t_ exceeding 1 indicate that the epidemic will tend to grow, while values below 1 indicate that the epidemic will tend to decline. We used a Poisson transmission model to estimate the time-varying reproduction numbers from serial intervals and incidence over time.^10,11^ Time-varying estimates of reproduction numbers were made with a 7-day sliding window using the R package EpiEstim, accounting for imported cases and assuming a serial interval of 4.7 days and a standard deviation of 2.9 days.^12^

To measure changes in influenza transmissibility over time, we first calculated the “ILI+” proxy^13,14^ for influenza A(H1N1) during the 2019/20 winter by multiplying together the weekly ILI consultation rates with the weekly proportions of specimens positive for influenza A(H1N1), which was the predominant strain. The ILI+ proxy is a better correlate of the incidence of influenza virus infections in the community than either ILI rates alone or laboratory detection rates alone.^14^ We interpolated daily ILI+ proxy values from the weekly influenza ILI+ proxy values by using flexible cubic splines.^15^

Using the daily ILI+ proxy, we estimated daily transmissibility via the effective reproduction number, R_t_. We used a simple branching process model for epidemic spread to estimate the time varying intensity of transmission.^16^ We assumed the serial interval distribution for influenza followed a gamma distribution with a mean of 2.85 days and standard deviation of 0.93 days.^17^ We repeated these analyses for the daily influenza A(H1N1) hospitalization rates among children in two large local hospitals. We evaluated the changes in transmissibility by comparing the *R*_*t*_ values during two weeks before and after the start of the school closure (including Chinese New Year holidays) for the 2019/20 winter influenza season. We compared the reductions in 2019/20 with reductions in previous years when the Chinese New Year holidays occurred during influenza epidemics. All analyses were conducted with R version 3.6.2 (R Foundation for Statistical Computing, Vienna, Austria).

## RESULTS

As of 11 March 2020, Hong Kong has confirmed 129 cases of SARS-CoV-2 infection, including 39 cases in persons that were presumed to have acquired infection outside of Hong Kong (“imported infections”), 30 cases that could not be linked to any other case (“unlinked infections”) and 60 cases that were linked to the other known cases (Figure 1). Among these 129 cases of SARS-CoV-2 infection, 15 were asymptomatic infections, while 114 were confirmed COVID-19 cases. Figure 1 shows the timeline of interventions that were implemented by the government in Hong Kong (Figure 1), including border restrictions, flexible working arrangements, and school closures from kindergartens up to tertiary and post tertiary institutions and including tutorial centres. By mid-February, some schools in Hong Kong had resumed teaching activities via the internet. Some religious organizations have cancelled services since February 13, and many conferences and other local mass gatherings have been cancelled. Quarantine orders have been issued to close contacts of confirmed cases, as well as travellers arriving from affected countries including mainland China, South Korea, Italy and Iran, and affected regions in France, Germany and Spain. To date, a total of 24,125 (20,516 residents and 3,609 non-residents) have undergone quarantine at several locations including at home, at hotels, and at designated quarantine facilities.^18^

**Figure 1.**
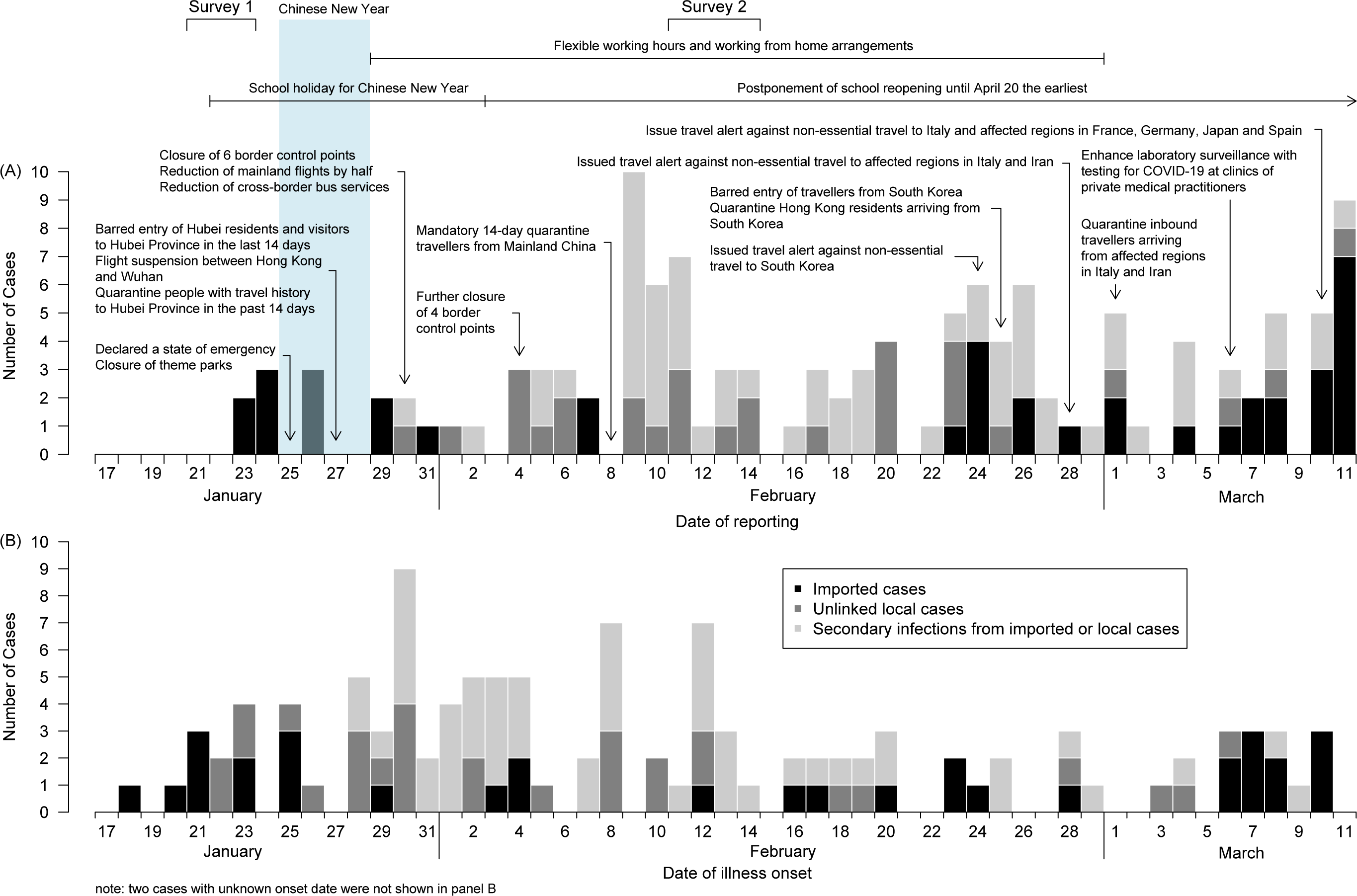
Detection of COVID-19 cases in Hong Kong by reporting date (panel A) and by onset date (panel B), with information on control measures implemented, and dates of population surveys. The Chinese New Year, a major winter festival in Hong Kong, began on Saturday January 25 and there were public holidays on January 25-28. Most schools started holidays on January 22 and were scheduled to resume on February 3. Due to the detection of COVID-19 cases in Hong Kong, the government deferred class resumption on January 27, January 31, February 13 and February 25 to February 16, March 2, March 16 and then April 20 at the earliest, respectively.

While unlinked COVID-19 cases as well as a number of clusters of cases have been detected, indicating that limited local transmission has been occurring, there is no evidence of exponential rise in the incidence of unlinked cases over time. Figure 2A shows the incidence of those unlinked cases. The estimated daily effective reproductive number which has remained below 1 throughout February and early March (Figure 2B). Data on influenza activity based on the community ILI+ proxy were very consistent with the rate of hospitalizations in children in Hong Kong (Figure 3A and 3B). Influenza activity peaked in the second week of January, with influenza A(H1N1) predominating, and declined to low levels by the second week of February. The effective reproductive number gradually declined from the second week of January to below 1 before Chinese New Year, rebounded to above 1 around Chinese New Year, and then declined again in early February (Figure 3C and 3D). The estimated R^t^ was 1.28 (95% confidence interval, CI: 1.26–1.30) before the start of the school holidays/closure and 0.72 (95% CI: 0.70– 0.74) during the holiday/closure weeks, corresponding to a 44% (95% CI: 34%–53%) reduction in transmissibility (Figure 3C). Similarly, the R_t_, calculated from hospitalization data dropped significantly after the start of the school holiday/closure by 33% (95% CI: 24%–43%) (Figure 2D). In comparison, we estimated the reduction in R_t_ to be a maximum of 15% (95% CI: 11%–19%) during the 2010/11 winter and 14% (95% CI: 7%–22%) during the 2014/15 winter due to the Chinese New Year holidays alone (Figure 4).

**Figure 2.**
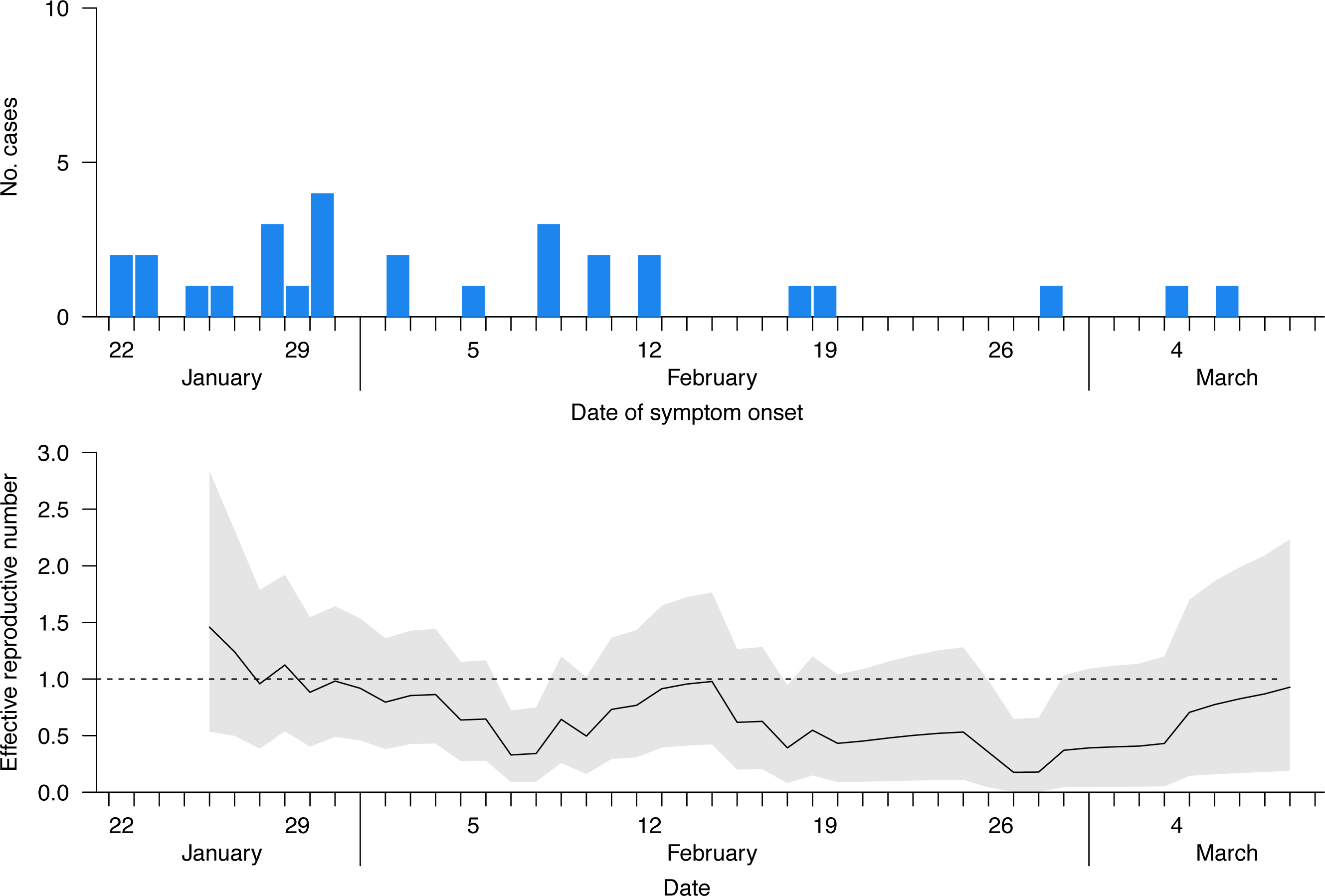
Panel A: Incidence of unlinked cases of COVID-19 in Hong Kong (blue bars). Panel B: Estimates of the daily effective reproductive number *R*_*t*_ of COVID-19 over time, with the gray shaded area indicating the 95% pointwise confidence intervals. The dotted line indicates the critical threshold of *R*_*t*_*=*1, if the reproductive number exceeds that for a prolonged period we would expect an epidemic to occur.

**Figure 3.**
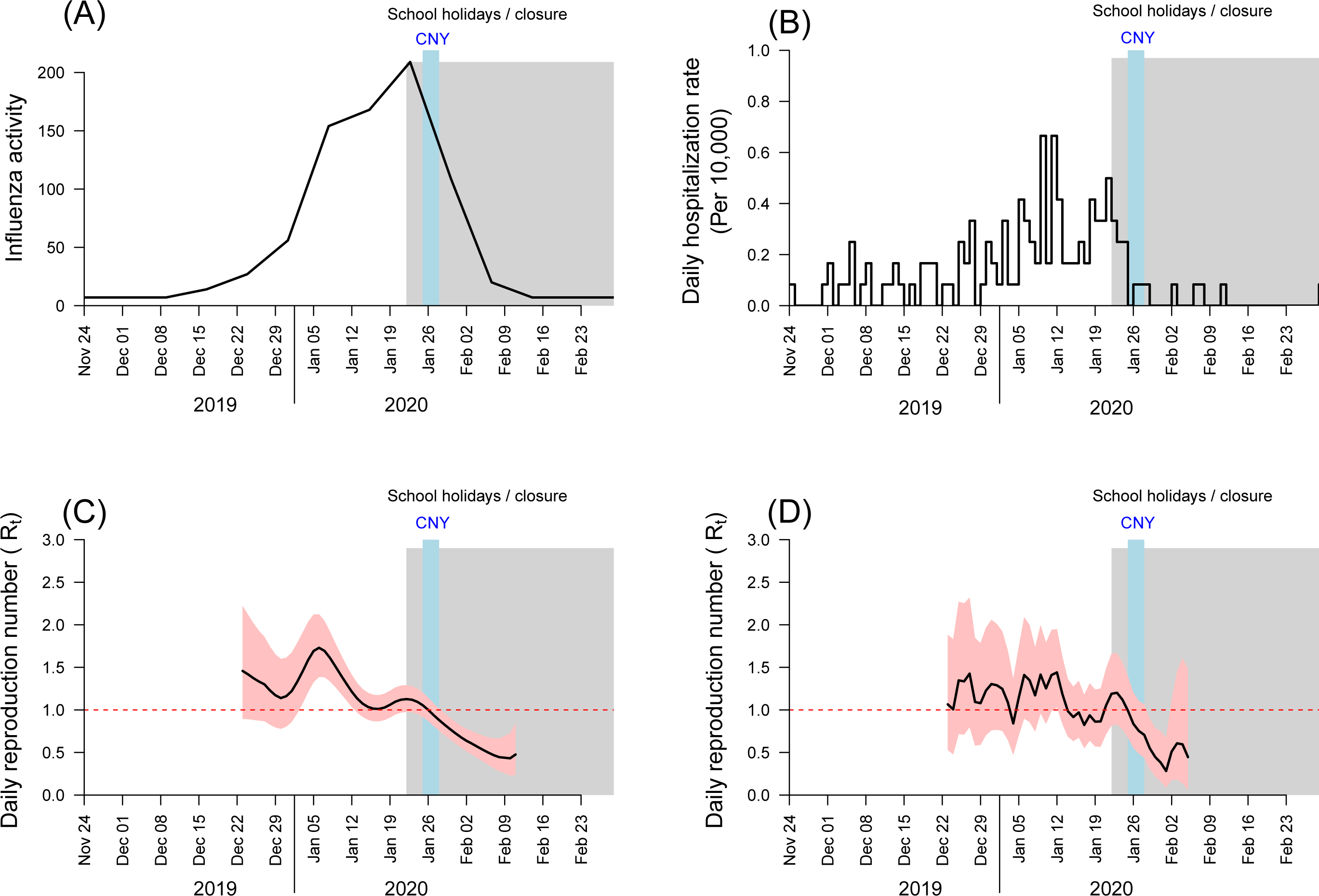
Panel A: weekly incidence rates of ILI+ for influenza A(H1N1), calculated as the weekly influenza-like illness rate multiplied by the proportion of laboratory specimens testing positive for influenza A(H1N1). Panel B: daily incidence rates of hospitalization with influenza A(H1N1) in children in two large hospitals in Hong Kong. Panel C: estimated effective reproductive number R_t_ in Hong Kong based on the ILI+ data, with 95% confidence interval indicated by the shaded region. Panel D: estimated effective reproductive number R_t_ in Hong Kong based on the hospitalization rate data, with 95% confidence interval indicated by the shaded region. Red dashed lines indicate transmission threshold (R_t_=1); Shaded bars show the dates of school holidays (Chinese New Year, in light blue) and school closure (in gray).

**Figure 4.**
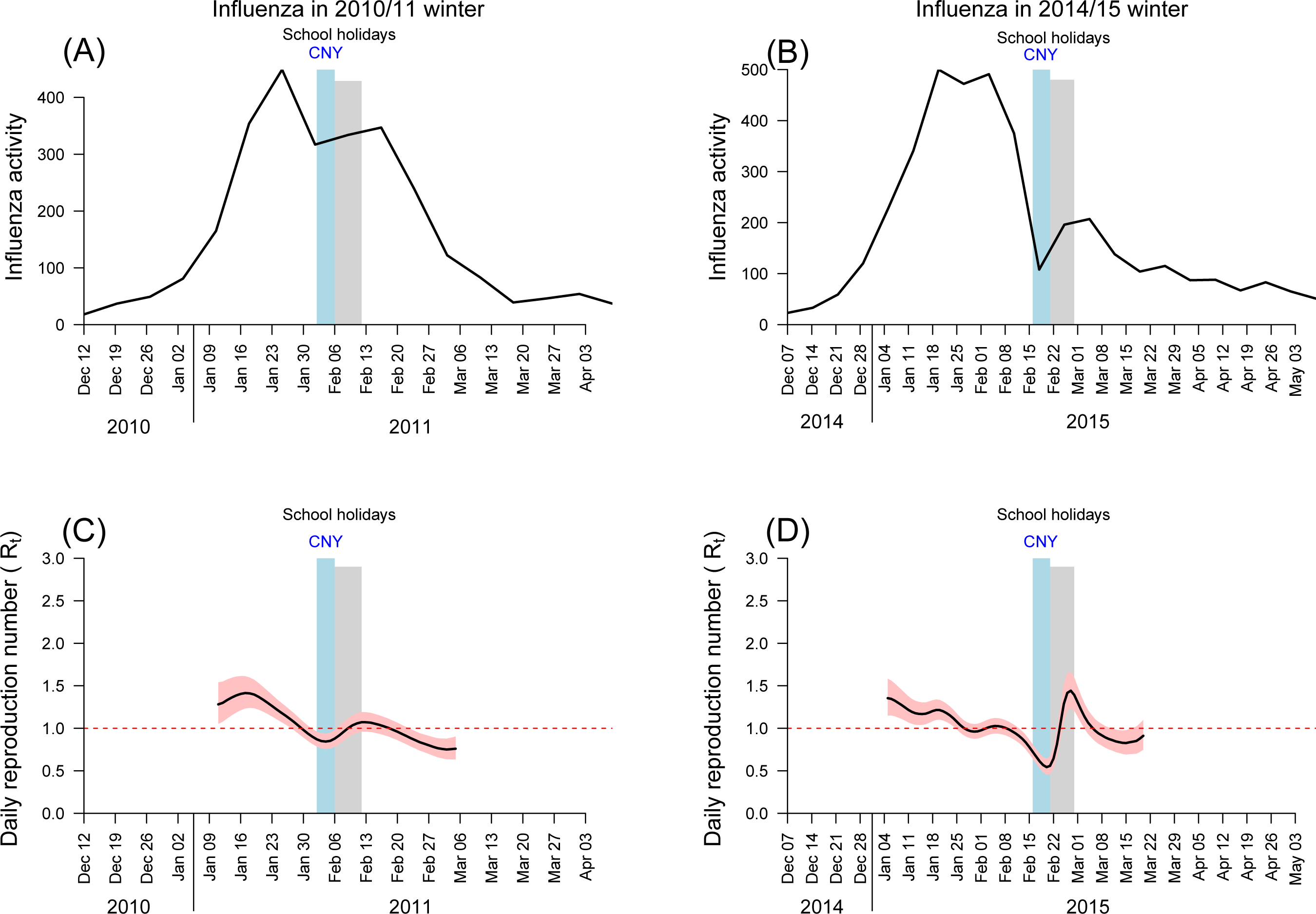
Weekly incidence rates of ILI+ for influenza (all type and subtypes), calculated as the weekly outpatient influenza-like illness rate multiplied by the proportion of laboratory specimens testing positive for influenza (A) for 2010/11 winter influenza season and (B) for 2014/15 winter influenza season. The estimated effective reproductive number R_t_ in Hong Kong based on the ILI+ data, with 95% confidence interval indicated by the shaded region (C) for 2010/11 winter season and (D) for 2014/15 winter season. Red dashed lines indicate transmission threshold (R_t_=1); Shaded bars show the dates of school holidays (in gray) and Chinese New Year timing (in light blue).

In our two surveys, we interviewed a total of 1008 and 1000 participants, respectively, with response rates among eligible respondents of 22% and 23% (Appendix Figure 1). Respondents included a broad cross-section of the adult population of Hong Kong (Table 1). Respondents perceived that they had similar susceptibility to COVID-19 as seasonal influenza, but that COVID-19 was much more serious infection, and around 80% of respondents reported worrying about being infected with COVID-19 (Table 2). In survey 2, 76% of respondents agreed with the statement that complete border closure would be effective in preventing the spread of COVID-19 to Hong Kong, 84.1% were worried about medical supplies including face masks, but only 27.7% were worried about the living supplies in Hong Kong (Table 2).

**Table 1.** Characteristics of respondents in two telephone surveys in Hong Kong.

| Characteristics | Survey 1<br>(20-23 January)<br>(N=1008) | Survey 2<br>(11-14 February)<br>(N=1000) |
| --- | --- | --- |
|  | n (%) | n (%) |
| **Sex** |  |  |
| Male | 472 (46.8) | 431 (43.1) |
| Female | 536 (53.2) | 569 (56.9) |
| **Age group, years <sup>†</sup>** |  |  |
| 18-24 | 99 (9.8) | 92 (9.2) |
| 25-34 | 170 (16.9) | 209 (20.9) |
| 35-44 | 189 (18.8) | 176 (17.6) |
| 45-54 | 162 (16.1) | 181 (18.1) |
| 55-64 | 172 (17.1) | 165 (16.5) |
| 65 or above | 202 (20.0) | 163 (16.3) |
| **Education level <sup>†</sup>** |  |  |
| Not educated/ pre-elementary | 15 (1.5) | 17 (1.7) |
| Primary | 82 (8.1) | 59 (5.9) |
| Secondary | 386 (38.3) | 387 (38.7) |
| Tertiary | 521 (51.7) | 531 (53.1) |
| **Occupation <sup>†</sup>** |  |  |
| Executive and professional | 281 (27.9) | 231 (23.1) |
| Clerical and service worker | 251 (24.9) | 258 (25.8) |
| Production worker | 73 (7.2) | 61 (6.1) |
| Student | 63 (6.2) | 54 (5.4) |
| Homemaker/ housewife | 126 (12.5) | 139 (13.9) |
| Retired person | 185 (18.4) | 173 (17.3) |
| Unemployed or not working for other reason | 15 (1.5) | 31 (3.1) |
| Others | 1 (0.1) | 42 (4.2) |

<sup>†</sup> 14 (1.4%) respondents did not report their age in both surveys, 4 (0.4%) and 6 (0.6%) respondents did not report their education level in survey 1 and 2, 13 (1.3%) and 11 (1.1%) respondents did not report their occupation in survey 1 and 2.

**Table 2.** Public attitudes, risk perceptions and behavioural responses towards COVID-19^a^ and seasonal influenza in two telephone surveys in Hong Kong.

| Items | Survey 1<br>(21-23 January)<br>(n=1008)<br>% (95% CI)<sup>b</sup> | Survey 2<br>(11-14 February)<br>(n=1000)<br>% (95% CI)<sup>b</sup> |
| --- | --- | --- |
| **Risk perception of COVID-19** |  |  |
| Perceived susceptibility to COVID-19 ("How likely do you think it is that you will contact COVID-19 over the next one month?" -- likely/very likely/ certain rather than never/very unlikely/unlikely/evens) | 18.9 (16.0, 21.9) | 17.4 (14.8, 20.1) |
| Perceived severity of COVID-19 (serious/ very serious rather than very mild/mild/moderate) | 89.6 (85.8, 93.3) | 90.5 (86.4, 94.7) |
| Worry about being infected with COVID-19 (moderately/ very much worried rather than not at all worried or slightly worried) | 52.5 (48.7, 56.3) | 53.9 (49.9, 57.9) |
| **Risk perception of seasonal influenza** |  |  |
| Perceived susceptibility to seasonal influenza ("How likely do you think it is that you will contact seasonal influenza over the next one month?" -- likely/ very likely/ certain rather than never/very unlikely/unlikely/evens) | 25.1 (22.0, 28.3) | 22.5 (19.4, 25.6) |
| Perceived severity of seasonal influenza (serious/ very serious rather than very mild/mild/moderate) | 42.3 (38.4, 46.3) | 32.7 (28.9, 36.6) |
| Worry about being infected with seasonal influenza (moderately/ very much worried rather than not at all worried or slightly worried) | 36.5 (32.9, 40.0) | 30.3 (26.6, 33.9) |
| **Attitudes towards COVID-19 (agree/ strongly agree to statements)** |  |  |
| I'm confident that I can take measures to protect myself against COVID-19 | 50.5 (46.6, 54.4) | 59.2 (54.9, 63.5) |
| I believe that the Hong Kong government can take effective measures to control the spread of COVID-19 in Hong Kong | 33.5 (29.9, 37.1) | 31.8 (27.8, 35.8) |
| I believe that the Central Chinese government can take effective measures to control COVID-19 | 31.7 (28.1, 35.3) | 39.0 (34.8, 43.2) |
| I believe that complete border closure is an effective measure to prevent COVID-19 from spreading from mainland China to Hong Kong | -<sup>c</sup> | 76.4 (72.3, 80.5) |
| Complete border closure will seriously affect the life of citizens | -<sup>c</sup> | 33.2 (29.2, 37.1) |
| I worry about the medical supplies such as facemask in Hong Kong | -<sup>c</sup> | 84.1 (80.0, 88.2) |
| I worry about the living supplies in Hong Kong due to border closure | -<sup>c</sup> | 27.7 (24.0, 31.4) |
| **Preventive measures taken against COVID-19** |  |  |
| In the past 7 days, did you take the following measure to prevent yourself from contracting COVID-19? |  |  |
| Avoid going to crowded places | 61.3 (57.2, 65.4) | 90.2 (86.2, 94.2) |
| Avoid visiting mainland China | 78.1 (73.9, 82.2) | -<sup>c</sup> |
| Avoid contact with people with respiratory symptoms | 66.8 (62.7, 70.9) | 80.0 (76.0, 84.0) |
| Use facemasks | 74.5 (70.4, 78.6) | 97.5 (93.5, 100) |
| Wash hands more often (including using hand sanitizer) | 71.1 (67.0, 75.2) | 92.5 (88.6, 96.5) |
| Avoid touching public objects or use protective measures when touching public objects (e.g. use tissue) | 36.4 (32.3, 40.5) | 73.8 (69.8, 77.9) |
| House disinfection | -<sup>c</sup> | 89.3 (85.2, 93.4) |
| Using serving utensils when eating | -<sup>c</sup> | 66.0 (61.9, 70.1) |
| Stay at home as much as possible | -<sup>c</sup> | 88.0 (83.9, 92.1) |
| Avoid going to health care facilities | -<sup>c</sup> | 81.0 (77.0, 85.1) |

<sup>a</sup> In the interviews, “武漢肺炎” (Wuhan pneumonia) was used because this term was most frequently used in the media while surveys were conducted
<sup>b</sup> Proportions were weighted by age and sex to the adult population in Hong Kong
<sup>c</sup> Questions not asked
Abbreviation: CI = confidence interval

We also identified considerable increases in the use of preventive measures in response to the threat of COVID-19. In recent years, face mask use in the general community has generally been restricted to individuals who are ill and those who feel particularly vulnerable to infection and want to protect themselves. In our first and second surveys we estimated that 74.5% and 97.5% of the general adult population wore masks when going out, 61.3% and 90.2% avoided going to crowded places, and 71.1% and 92.5% reported washing or sanitizing their hands more frequently, respectively (Table 2). In survey 2, 88.0% reported staying at home as much as possible.

In our second survey which was conducted from Tuesday 11 February through Friday 14 February, we also asked the subset of respondents who were parents of school-age children about their support for school closures and the activities of their children during the school closures. Among respondents who were parents, 249/261 (95.4%) agreed or strongly agreed that school closure is needed as a control measure for COVID-19 in Hong Kong, and 207/261 (79.3%) responded that their children had no contact with persons other than their household members on the preceding day. More detailed information on behaviours of children during the school closures will be reported separately.

## DISCUSSION

Our findings suggest that the package of non-pharmaceutical interventions that Hong Kong has implemented since the latter part of January 2020 have succeeded at containing spread of COVID-19 with little evidence indicating sustained local spread beyond sporadic cases and known clusters, seven weeks (also about six generation times) since the first known case showed symptoms in Hong Kong. If our findings are borne out elsewhere, they support the perspective that COVID-19 can be meaningfully controlled, or at least mitigated, by familiar social distancing and population behavioural changes short of the draconian measures introduced in mainland Chinese cities.

It would however be premature to conclude at this early date that COVID-19 is being completely contained. However, our findings here indicate that control measures and changes in population behaviour have led to a substantial reduction in influenza transmission in early February 2020 (Figure 3). While it is not possible to distinguish the specific effects of each community measure, it is clear from Figure 1 that the travel restrictions have reduced the number of imported cases, with very few detected during February, and only a resurgence in early March with imported cases from Europe and the middle East.

Medical isolation of cases and intensive contact tracing and strict quarantine should have limited onwards transmission from locally confirmed cases and their contacts. However, it is likely that some infected persons have not been identified. Figure 1 shows the onset dates of 30 unlinked cases, i.e. cases for whom the infector could not be determined. If there were an increase over time in unlinked cases, it would indicate that incidence in the community could be increasing through silent transmission. However, our analyses suggest that reductions in population mixing have substantially reduced influenza transmission (Figure 3). If COVID-19 transmission occurs through similar modes and routes to influenza virus transmission, we would also expect substantial reductions in COVID-19 transmissibility, although a potentially higher basic reproductive number for COVID-19 indicates that it might be more difficult to control than influenza.^19^

The estimated 44% reduction in transmission in the general community in February 2020 (Figure 3C) was much greater than the estimated 10% to 15% reduction in transmission conferred by school closures alone during the 2009 pandemic,^20^ and the 16% reduction in transmission of influenza B conferred by school closures during the 2017/18 winter in Hong Kong.^21^ We therefore conclude that the other social distancing measures and avoidance behaviours have had a substantial effect on influenza transmission on top of the effect of school closures. However, we note that if the basic reproductive number of COVID-19 in Hong Kong exceeds 2, noting that it was 2.2 in Wuhan,^19^ we would need more than a 44% reduction in COVID-19 transmission to completely avert a local epidemic. A reduction of this magnitude could, however, substantially flatten the peak and area under the epidemic curve, thus reducing the risk of exceeding the capacity of the healthcare system, and potentially saving hundreds or even thousands of lives especially amongst older adults.

The postponement of class resumption in local schools in Hong Kong is strictly speaking a class dismissal or suspension rather than a school closure, because most teachers are still required to go to school premises to plan e-learning activities and set homework. School closures have been implemented locally in previous years, including during the SARS epidemic in 2003,^6^, during the influenza pandemic in 2009,^20^ and to control seasonal influenza epidemics in 2008 and 2018.^21,22^ While school closures can have considerable effects on influenza transmission, their role in reducing COVID-19 transmission would depend on the susceptibility of children to infection and their infectiousness if infected. Both of these are major unanswered questions at present.^23,24^ Despite this acknowledged uncertainty, our survey revealed considerable local support for school closures at present.

The general community in Hong Kong have also changed their individual behaviours in response to the threat of COVID-19. People have been choosing to stay at home more, and in our most recent survey more than 90% of respondents reported to avoid going to crowded place and 98% of them wearing face masks when leaving home (Table 2). Using similar surveys, face mask use during SARS in 2003 was 79%,^5^ but at most 10% during the influenza A(H1N1) pandemic in 2009.^7^ These major changes in behaviour indicate the level of concern among the local population for this particular infection, and the level of voluntary social distancing in addition to the distancing created by school closures.

There are some limitations of our study. First, we cannot identify whether containment measures, social distancing measures or behavioural changes are most important in suppressing COVID-19 transmission. It is likely that each plays a role, noting that unlinked cases have been identified in the community and will continue to be identified, indicating that not every chain of transmission has been identified by contact tracing from known cases. While we have demonstrated major effects of control measures and behavioural changes on influenza transmission (Figure 3), we can only infer similar changes in COVID-19 transmissibility if the two viruses share similar dynamics of transmission. Second, our survey of population behaviours could have been affected by response bias, since we relied on self-reported data, and could have been affected by selection bias away from working adults, although this should have been reduced by conducting surveys in non-working as well as working hours. Without data on the non-respondents, we were unable to assess potential selection bias. Without a baseline survey before 23 January we could not compare changes in behaviours, although we have published the results of similar surveys from previous epidemics which can be used for comparison.^6-8,19^ Finally, while we identified reductions in the incidence of influenza virus infections in outpatients and paediatric inpatients (Figure 3), it is possible that these time series were affected by reduced health-care seeking behaviours and limited healthcare access that probably resulted from private clinic closure, which occurred around the period of the Chinese New Year holiday.

In conclusion, measures taken to control the spread of COVID-19 have been effective and have also had a substantial impact on influenza transmission in Hong Kong. While the transmission dynamics and modes of transmission of COVID-19 have not been precisely elucidated, they are likely to share at least some characteristics with influenza virus transmission, as both are directly transmissible respiratory pathogens with similar viral shedding dynamics.^25^ The measures implemented in Hong Kong are less drastic than those used to contain transmission in Wuhan in the past month, but are probably more feasible in most locations. If these measures and population responses can be sustained, avoiding fatigue among the general community, they should meaningfully mitigate the impact of a local epidemic of COVID-19.

## Data Availability

Data are available from the first author on request

## ACKNOWLEDGMENTS

The authors thank Julie Au for administrative support

## FIGURE LEGENDS

Appendix Figure 1. Flow chart of calls made, calls answered, and respondents successfully interviewed in two telephone surveys in Hong Kong, on 20-23 January and 11-14 February, respectively.

